# Aberrant levels of cortical myelin distinguish individuals with unipolar depression from healthy controls

**DOI:** 10.1101/2021.02.25.21252472

**Authors:** David A.A. Baranger, Yaroslav O. Halchenko, Skye Satz, Rachel Ragozzino, Satish Iyengar, Holly A. Swartz, Anna Manelis

## Abstract

The association of unipolar depression (UD), relative to healthy controls (HC), with cortical myelin is underexplored, despite growing evidence of associations with white matter tract integrity. We characterized cortical myelin in the 360 Glasser atlas regions using the T1w/T2w ratio in 39 UD and 47 HC participants (ages=19-44, 75% female). A logistic elastic net regularized regression with nested cross-validation and a subsequent linear discriminant analysis conducted on held-out samples were used to select brain regions and classify UD vs. HC. True-label model performance was compared against permuted-label model performance. Cortical myelin distinguished UD from HC with 68% accuracy (p<0.001; sensitivity=63.8%, specificity=71.5%). Brain regions contributing to this classification performance were located in the orbitofrontal cortex, anterior cingulate, extended visual, and auditory cortices, and showed statistically significant decreases and increases in myelin levels in UD vs. HC. The patterns of cortical myelin in these regions may be a biomarker of UD.

## 1 INTRODUCTION

Unipolar depression (UD) is a leading cause of disability worldwide (1), with an economic burden of $210 billion dollars in the United States alone (2). Despite its impact, treatments for the disorder remain ineffective for many patients (3). Thus, there is a pressing need to understand the neurobiological etiology of UD to facilitate the development of improved treatments and prevention strategies.

Unipolar depression is characterized by dysfunctional affective and cognitive processing (4), including reduced executive functioning (5), biased emotional processing (6), and impaired reward processing (7). Correspondingly, individuals with UD show aberrant activation during tasks which recruit these processes, including activation in the striatum, hippocampus, amygdala, orbitofrontal cortex, prefrontal cortex, insula, cingulate, and occipital cortex (8–11). In addition, a growing body of literature has reported structural abnormalities associated with depression in many of these same regions described above, both in grey matter (12–14) and in white matter (15,16). Meta-analyses of diffusion weighted imaging (DWI) studies have repeatedly found evidence for lower fractional anisotropy (FA) in depressed populations (17–19). More recently, studies using very large samples (i.e., the UK Biobank), as well as meta-analyses combining both published and unpublished data (i.e., the ENIGMA consortium), have observed widespread and replicable reductions in FA(15,16). Notably, white matter integrity in identified regions has been shown to correlate with the cognitive processes disrupted in depression, including processing speed (20,21), emotion regulation (22,23), and reward learning (24).

Emerging evidence suggests that cortical myelin may be impacted in individuals with UD and that it may partially mediate some of the cognitive processes that are impaired in affected individuals. For example, studies of *post mortem* brain tissue from donors with UD have observed reduced myelination, a reduction in the number of oligodendrocytes and other glia (cells whose functions include generating and maintaining myelin (25)), and reduced expression of oligodendrocyte lineage genes (26,27). A study of individuals with treatment resistant depression revealed a reduced magnetization transfer ratio (MTR), which is thought to reflect lower myelin levels, in the cingulate cortex and insula (28). A recent study using R1 (1/T1) as a measure of myelination observed reduced whole-brain myelin, but no significant difference in cortical myelin in a handful of *a priori* bilateral regions between individuals with depression and healthy controls (HC) (29),

Developments in magnetic resonance imaging (MRI) methodology permit the examination of cortical myelin via the T1w/T2w ratio (30,31). Studies in population-based samples using this metric have found that lower myelin in the cingulate, orbitofrontal cortex, and middle temporal cortex correlated with poor sleep quality (32), lower frontal-pole myelin and greater myelin in the occipital cortex correlated with neuroticism (33), and lower myelin in the motor and higher myelin in the insular, cingulate, prefrontal, and superior parietal cortices correlated with trait anxiety (34). While poor sleep, high neuroticism, and trait anxiety might represent concurrent symptoms of depression, prior studies have not systematically examined cortical myelin in participants with depression as compared to HC.

The goals of the present study were (1) to ascertain whether cortical myelin content (characterized by the T1w/T2w ratio) is predictive of unipolar depression (UD), and (2) to characterize the brain regions that are predictive of case/control status. Based on the prior studies mentioned above, we hypothesized that cortical myelin levels would distinguish individuals with UD from HC and that these differences will be especially pronounced in the prefrontal cortical (PFC), cingulate, parietal and occipital regions that support reward and emotional processing, which are dysregulated in UD (11,35).

## 2 METHODS

### 2.1 Participants

The study was approved by the University of Pittsburgh Institutional Review Board. Participants were recruited from the community, universities, and counseling and medical centers. They gave written informed consent, were right-handed, fluent in English, and were matched on age and sex. Individuals with unipolar depression (UD) met DSM-5 criteria for major depressive or persistent depressive disorders. Healthy controls (HC) had no personal or family history of psychiatric disorders. Exclusion criteria included a history of head injury, metal in the body, pregnancy, claustrophobia, neurodevelopmental disorders, systemic medical illness, premorbid IQ<85 per the National Adult Reading Test (36), current alcohol/drug abuse, Young Mania Rating Scale scores>10 (YMRS(37)) at scan, or meeting criteria for any psychotic-spectrum disorder. Data were drawn from an ongoing longitudinal study that includes neuroimaging sessions at baseline and 6-month follow-up and clinical evaluations at baseline, 6-months, and 12-months. The present report includes baseline data available for 55 HC and 50 UD. Participants were excluded from analyses due to (1) previously undetected brain abnormalities of potential clinical relevance: 2 UD, (2) diagnosis conversion during the course of the study: 1 HC was diagnosed with major depressive disorder, and 1 UD was diagnosed with bipolar disorder; (3) scanner or movement-related artifacts in MRI data (4 HC, 7 UD), and (4) poor-quality myelin maps (see Section 2.4.2.1 *Subject-level processing;* 3 HC, 1 UD). The final sample included 47 HC and 39 UD.

### 2.2. Clinical assessment

All diagnoses were made by a trained clinician and confirmed by a psychiatrist according to DSM-5 criteria using SCID-5 (38). Additional information collected included illness onset and duration, number of current episodes, comorbid psychiatric disorders, current depression symptoms using the Hamilton Rating Scale for Depression (HRSD-25) (39), current mania symptoms using the Young Mania Rating Scale (YMRS) (37), and lifetime depression and hypo/mania spectrum symptomatology using the Mood Spectrum Self Report (MOODS-SR) (40). A total psychotropic medication load was calculated for each participant with UD, with greater numbers and doses of medications corresponding to a greater medication load (41,42).

### 2.3 Neuroimaging data acquisition

The neuroimaging data were collected at the University of Pittsburgh/UPMC Magnetic Resonance Research Center using a 3T Siemens Prisma scanner with a 64-channel receiver head coil and named according to the ReproIn convention (43). The DICOM images were converted to BIDS dataset using *heudiconv* (44) and *dcm2niix* (45). High-resolution T1w images were collected using the MPRAGE sequence with TR=2400ms, resolution=0.8×0.8×0.8mm, 208 slices, FOV=256, TE=2.22ms, flip angle=8°. High-resolution T2w images were collected using TR=3200ms, resolution=0.8×0.8×0.8mm, 208 slices, FOV=256, TE=563ms. Field maps were collected in the AP and PA directions using the spin echo sequence (TR=8000, resolution=2×2×2mm, FOV=210, TE=66ms, flip angle=90°, 72 slices).

### 2.4 Data analyses

#### 2.4.1 Clinical data analysis

HC and UD groups were compared on demographic and clinical variables using t-tests and chi-square tests. All analyses were conducted in R (https://www.r-project.org/).

#### 2.4.2 Neuroimaging data processing

##### 2.4.2.1 Subject-level preprocessing

Data quality was examined using *mriqc version 0*.*15*.*1* (46) and visually inspected (Supplemental Methods). Each participant’s cortical myelin was characterized with the T1w/T2w ratio (30,47,48) using the PreFreeSurfer, FreeSurfer, and PostFreeSurfer minimal preprocessing pipelines for the human connectome project (47). *Workbench v1*.*4*.*2* and *HCPpipelines-4*.*1*.*3* were installed system-wide on a workstation with GNU/Linux Debian 10 operating system. The spin echo field maps collected in AP and PA phase encoding directions were used for bias field correction in *PreFreeSurfer*. Registration to standard space was achieved via MSMSulc (49) in *PostFreeSurfer*. If FreeSurfer images and myelin maps had artifacts and gross errors (e.g., large regions of apparent low myelin in the occipital cortex due to the transverse sinus interfering with accurate identification of the pial surface), the data were removed from analyses. The resulting myelin maps were parcellated in *Workbench* using the 360 region Glasser Atlas (48). Further data quality assurance resulted in the removal of 11 outlier parcels (Supplemental Methods), leaving 349 parcels.

#### 2.4.3 Neuroimaging data analysis

##### 2.4.3.1 Elastic net and linear discriminant analysis

Neuroimaging data decoding studies can capitalize on complex relationships between variables, but their large numbers can present a challenge in deriving a generalizable model. The elastic net approach has emerged as a flexible tool for use with neuroimaging data (50) as it is able to reduce the influence of overly large coefficients and reducing the number of variables while generating multivariate models predictive of complex behaviors (51–53). Elastic net is a regularized regression which combines lasso and ridge regression (i.e., L1- and L2-norm regularization) (54). Ridge regression penalizes overly large coefficients, while lasso regression removes variables with small coefficients.

We used logistic elastic net regularized regression (55) to select variables (brain parcels) that were most predictive of case/control status. Elastic net has two parameters: alpha (*α)* controls the balance between the ridge and lasso regularizations, and lambda (*λ)* controls the strength of regularization. To provide equal contribution of each penalty to the loss function, we used *α*=0.5. To avoid model overfitting and bias, we implemented nested cross-validation to identify the optimal *λ* parameter. A linear discriminant analysis (LDA) (56) model was subsequently trained using selected variables to make out-of-sample prediction on held-out participants. This strategy is illustrated in Figure 1A and described in detail in Supplemental Methods. For each repetition of the nested cross-validation loop, two participants (1 UD and 1 HC) were held out. The rest of the sample was used to identify the optimal *λ* parameter which were then used to fit the elastic net model and select variables whose myelin levels were predictive of UD/HC status. These variables were then used to train an LDA model, which was tested on held-out participants. Results (model fit, variable selection, prediction accuracy) were evaluated against results from a permuted-label analysis (Figure 1B and Supplemental Methods).

**Figure 1.**
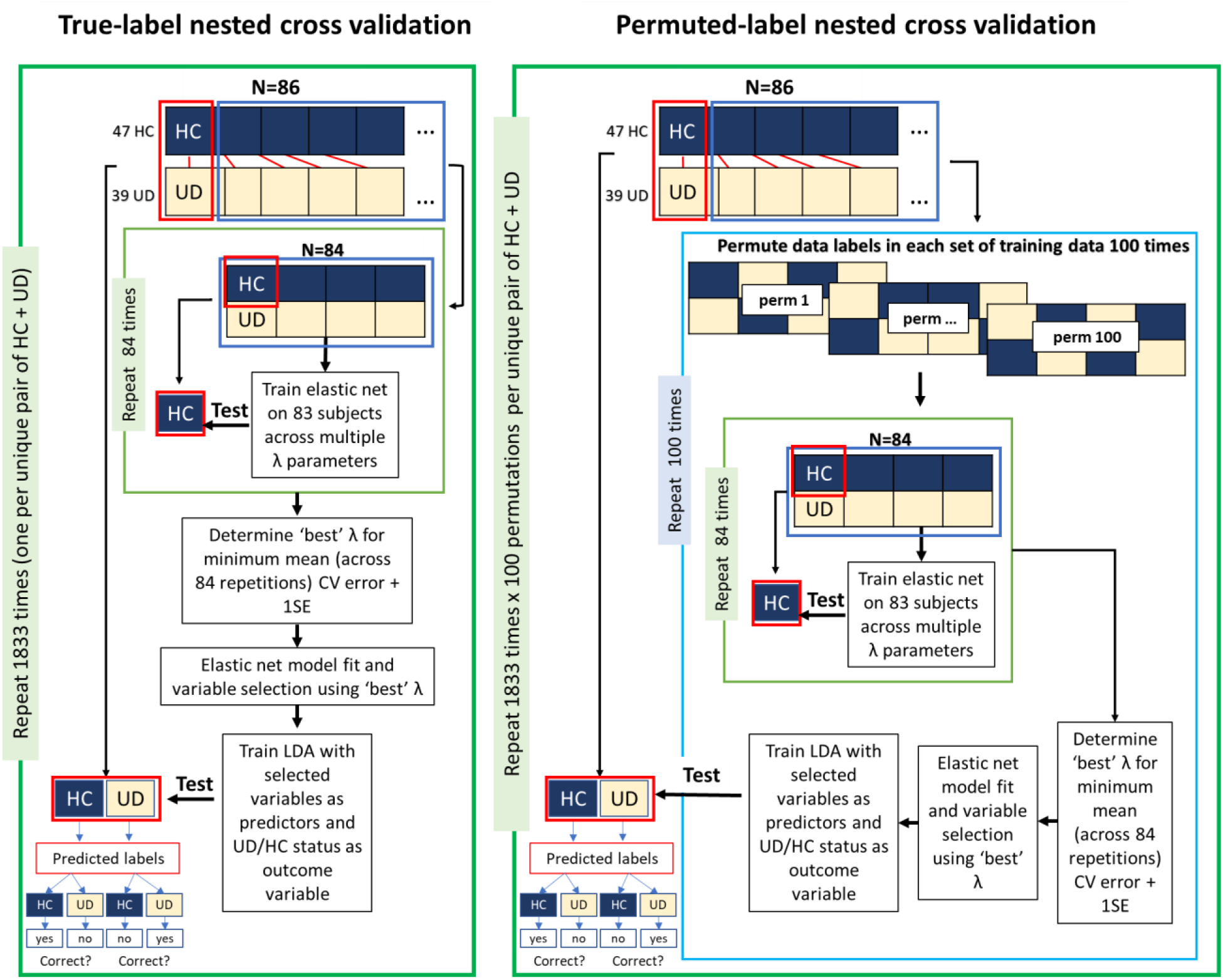
Diagram of analysis steps. Conceptual depiction of analysis steps including: (1) a unique pair of one UD and one HC participant is held-out; (2) an elastic net regression is used to select variables; (3) the retained variables are used an LDA model predicting case/control status; (4) the LDA model is tested on the held-out sample; (5) this process is repeated for each of the n=1833 pairs of subjects; (6) for each held-out pair, the training procedure is repeated with 100 unique permutations.

#### 2.4.3.2 Post-hoc analyses

##### 2.4.3.2.1 The relationship between true-label and permuted-label sample demographics

To ensure that the permuted results were not due to changes in the internal structure of the permuted samples in terms of demographic variables (i.e., age, sex and IQ), we compared the age, sex and IQ values in the permuted samples with that in the true-label samples.

##### 2.4.3.2.2 Association of LDA accuracy and cortical myelin with demographic and clinical characteristics

To further characterize the parcels selected by the logistic elastic net regression, we compared cortical myelin in UD vs. HC while controlling for age, sex, and IQ. To assess the potential influence of confounding variables on model fit, we tested whether demographic or clinical characteristics were predictive of classification accuracy. Regressions tested whether the joint effect of group, the variable in question, or their interaction, was associated with participant-wise accuracy (57). Additional models tested whether clinical characteristics of UD participants were associated with model accuracy. To assess the influence of confounding variables on variable selection, regressions similarly tested the association of demographic and clinical variables with cortical myelin, controlling for age, sex, and IQ. Results for each variable were separately corrected for multiple comparisons using false discovery rate (FDR).

#### 2.4.3.3 Exploratory analysis of a HC participant who was diagnosed with major depressive disorder 12 months from the baseline scan

One participant entered the study as a HC but was diagnosed with major depressive disorder sometime between 6 and 12 months after study onset. At the study visit at 12 months the participant had mild depressive symptoms. While this participant was excluded from all primary analyses described above, exploratory analyses investigated whether the myelin was predictive of the participant’s conversion from HC to UD. This analysis used the primary 86 participants and the variables selected in primary analyses (cortical myelin in 33 parcels and IQ) to train an LDA model. UD/HC status was then predicted at study onset and at the 6-month follow-up (both time points were prior to conversion).

## 3 Results

### 3.1 Sample demographics

Individuals with UD did not differ by age or sex but had higher IQ and current and lifetime depression severity compared to HC (Table 1).

**Table 1.** Demographic and clinical characteristics

|  | HC (mean/sd or count/percent) | UD (mean/sd or count/percent) | t-test or chi-squared test<br>HC vs. UD |
| --- | --- | --- | --- |
| N | 47 (54.7%) | 39 (45.3%) |  |
| Gender (number females) | 36 (76.6%) | 29 (74.6%) | $\chi^2(1)=0$, $p=1$ |
| UD diagnoses (MDD/PDD) | na | 26/13 | na |
| Age (years) | 28.55 (6.15) | 29.07 (6.88) | $t(84)=-0.37$, $p=0.71$ |
| IQ (NART) | **106.72 (6.24)** | **110.2 (7.49)** | **$t(84)=-2.35$, $p=0.02$** |
| Illness Onset (year of age) | na | 15.05 (5.02) | na |
| Lifetime episodes of depression | na | 3.25 (1.5) | na |
| Current depression severity (HRSD-25) | **1.74 (2.16)** | **12.69 (6.77)** | **$t(84)=-10.48$, $p<.01$** |
| Lifetime depression (MOODS-SR) | **2.15 (2.27)** | **18.41 (4.36)** | **$t(84)=-22.21$, $p<.01$** |
| Number taking Antidepressants | na | 22 (56.4%) | na |
| Number taking Mood stabilizers | na | 3 (7.7%) | na |
| Number taking Antipsychotics | na | 1 (2.6%) | na |
| Number taking Benzodiazepines | na | 5 (12.8%) | na |
| Number taking Stimulants | na | 3 (7.7 %) | na |
| Mean number of psychotropic medications | na | 1.05 (1.12) | na |
| Mean total medication load | na | 1.33 (1.53) | na |
| Number with comorbid diagnoses | na | 27 (69.2%) | na |

Note: Comparison of demographic and clinical characteristics between healthy controls (HC) and individuals with Unipolar Depression (UD). Tests were run as t-tests or chi-squared tests, as specified. Bold = $p<0.05$ . na=not applicable.

### 3.2 UD vs. HC LDA nested cross-validation classification accuracy

Cortical myelin levels and IQ distinguished UD from HC in subjects held-out during cross-validation, with an average accuracy of 68% (Figure 2A; sensitivity (UD): 63.8%, specificity (HC): 71.5%). The mean participant-wise accuracy across 84 classification loops ranged from 0% to 100%, and 84% of participants were classified either quite accurately (>80% n=53) or inaccurately (<20% n=19). When demographic variables were excluded from the LDA classification, nested cross-validation achieved 69% accuracy (UD: 65.2%, HC: 73%). Notably, while the mean classification accuracy was higher in the HC group, this difference was not statistically significant (t=0.88, p=0.38).

**Figure 2.**
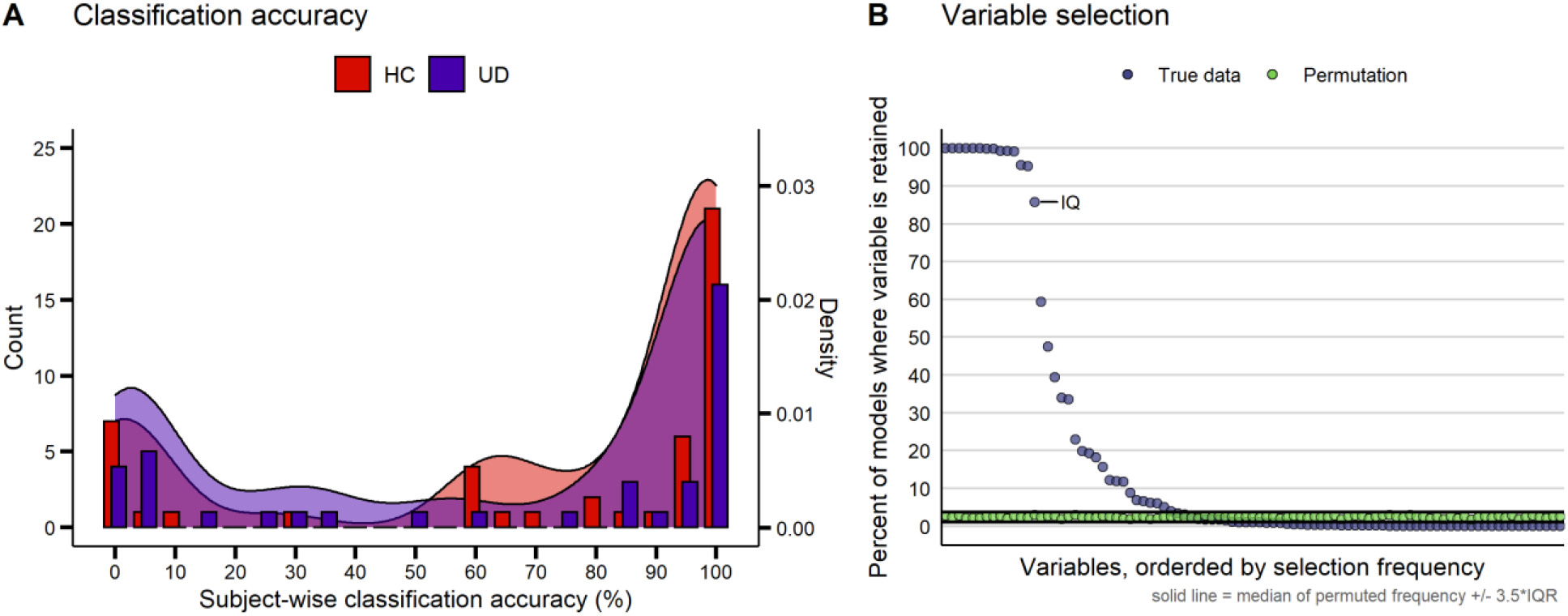
Classification accuracy and variable selection. A) Average subject-wise nested cross-validation classification accuracy in both healthy controls (HC) and participants with unipolar depression (UD). B) Variable selection frequency (percent of models where a given parcel was retained) in the true data (light blue) and permutations (green). The n=90 parcels which were retained at least once are shown. IQ, the only demographic variable selected, is labeled. Solid black lines represent the median of the variable selection frequency for all n=350 variables +/-3.5 x the interquartile range (IQR). See Supplemental Table 2 for the full list of variable selection frequencies.

In permutation analyses, no variable was selected in 66.9% of 183300 models. Within each participant, the proportion of models that did not select any variables ranged from 65.4% to 68.7%. Excluding instances when no variables were selected, the average participant-wise LDA accuracy in permutation analyses ranged from 41.2% to 59.2%, with an average accuracy of 50.5% (i.e., chance level). The test of whether age, sex and IQ in permuted samples differed from that in the sample with true labels showed that the demographics of the permuted groups differed (p<0.05, uncorrected) from the true-label groups only in 1.7% of cases.

**Table 2.**
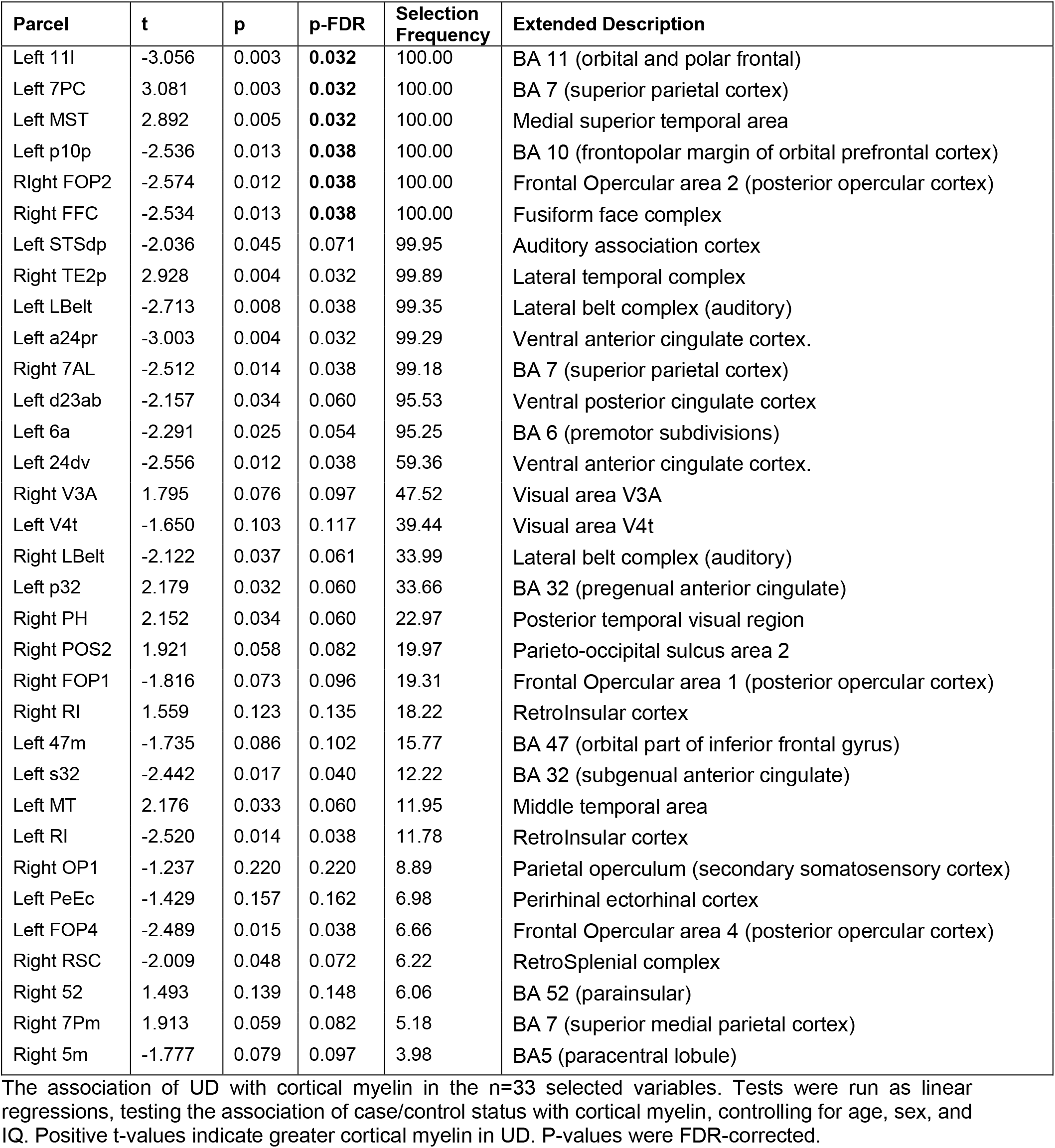
Association of unipolar depression with cortical myelin in selected parcels

### 3.3 Elastic net variable selection

*True-label* nested cross-validated elastic net models predicting diagnostic status (UD vs. HC) with 349 myelin parcels, age, sex and IQ, selected 90 myelin parcels (Supplemental Table 1) and IQ in at least one model (Figure 2B). Each model selected between 9 and 68 variables, with a median of 17 variables. *Permuted-label* nested cross-validated elastic net models selected all predictor variables at least once, but no variable was selected by more than 3% of the models. In addition, 66.9% of models with permuted labels did not select any variable at all. Within permuted-label models where at least one variable was selected, the number of selected variables ranged from 1 to 113, with a median of 17 variables selected.

Given that the variable selection frequency in the models with permuted labels likely represents noise, we applied the criterion of the median + 3.5*IQR across all permuted variables (3.77%) as the cutoff value to separate the potential noise variables from ‘signal’ variables in the nested cross-validated elastic net with true labels. This latter analysis identified 33 out of 90 myelin parcels, plus IQ, across the true-label nested cross-validated elastic net models that were above the cutoff line (Table 2, Figure 2B). These 33 parcels included multiple regions in the orbitofrontal cortex, insula, cingulate, and frontal operculum, as well as regions in the auditory and visual cortices (Supplemental Figure 3). No model retained the age and sex variables.

**Figure 3.**
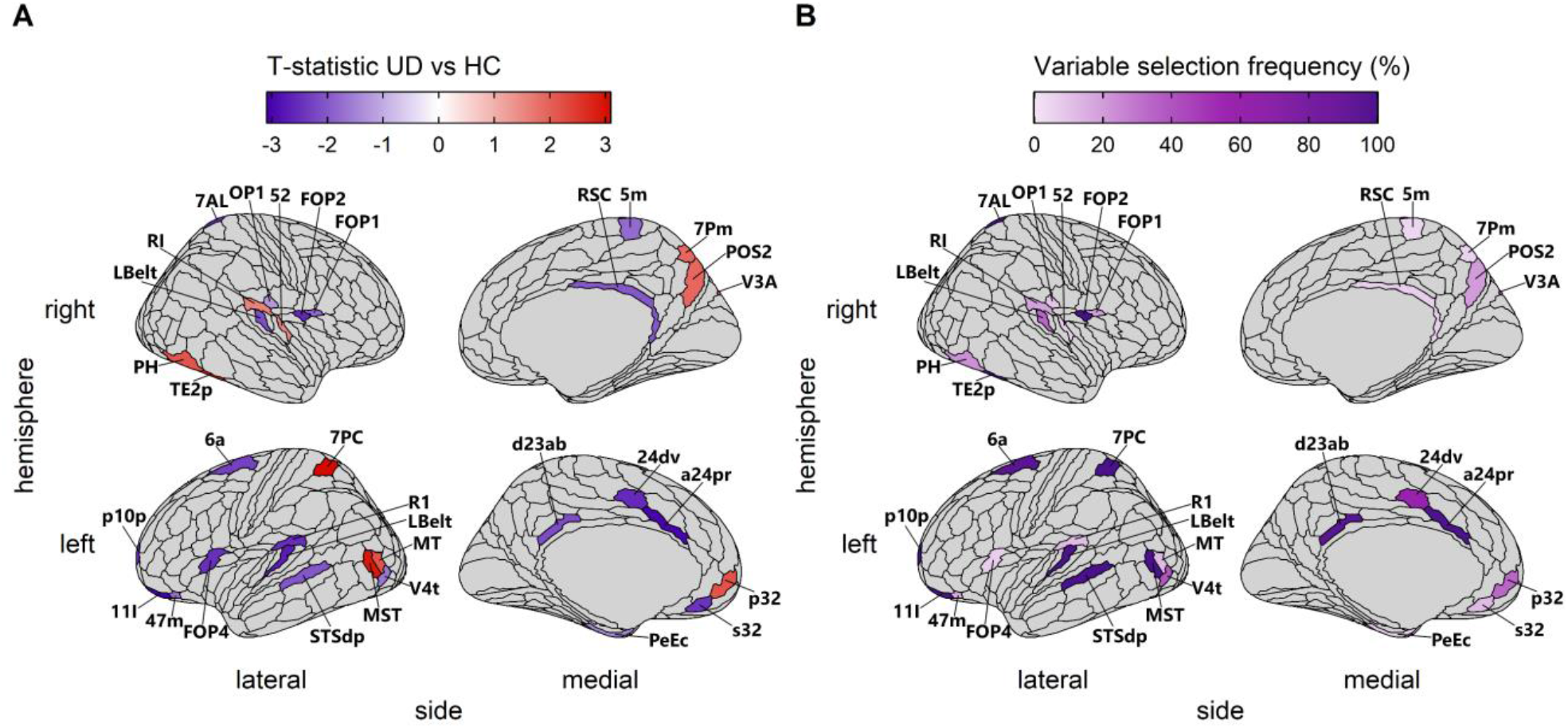
Association of diagnostic status with cortical myelin in selected parcels. A) The t-statistic for the association of unipolar depression (UD) with cortical myelin in the n=33 selected parcels. Regions where UD participants had greater average myelin than HC participants are in red, and regions where HC participants had greater average myelin than UD participants are in blue. B) The percent of models in which each parcel was retained. Regions are individually labeled (note that FFC is not visible, as it lies on the ventral surface).

Post-hoc analyses tested the association of diagnostic status with cortical myelin levels in the 33 selected myelin parcels (Table 2, Figure 3). These parcels include both regions where UD participants have lower mean cortical myelin than HC and regions where UD participants have greater mean cortical myelin, as well as regions where the two groups do not differ in their average level of cortical myelin. After FDR-correction for multiple comparisons, fourteen of the parcels showed evidence of significant differences between UD and HC participants (Table 2). An additional 8 parcels showed nominally significant differences between UD and HC participants (Table 2; p<0.05 uncorrected). Parcels that showed a greater absolute mean difference between the groups were selected more frequently in the nested cross-validation analysis (*r*=0.7, *p*=4×10^−6^).

The 33 selected parcels included 13 parcels in regions important for executive functioning and cognitive control, located in the anterior cingulate (left a24pr, left 24dv, left p32, and left s32), orbitofrontal cortex (left 11l, left p10p, and left 47m), posterior cingulate (right RSC, and left d23ab), frontal operculum (right FOP1 and left FOP4), and parietal cortex (right POS2 and right 7Pm). UD was largely associated with reduced cortical myelin in these regions, except in one region of the anterior cingulate (left p32), and the two regions in the parietal cortex (right POS2 and right 7Pm), where it was associated with increased myelin. An additional 19 of the identified parcels play roles in visual, somatomotor, and auditory processing. Visual processing regions included extrastriate regions (right V3A, left V4t, and left MT), ventral stream regions (right FFC and right TE2p), and dorsal stream regions (left MST, right PH, left 7PC, right 7AL). Auditory processing regions included regions in the auditory cortex (left LBelt, right LBelt, left RI, and right RI) and regions implicated in language (left STSdp, left FOP2, and right 52). Somatomotor regions included regions implicated in somatosensation (right 5m and right OP1) and a region in the premotor cortex (left 6a). UD was largely associated with reduced cortical myelin in auditory and somatomotor regions, except for two auditory regions (right 52 and right RI) where it was associated with increased myelin. In contrast, participants with UD had greater myelin in 6 visual regions and had reduced myelin in 3 regions (left V4t, left FFC, and left 7AL).

### 3.5 Post-hoc analyses

#### 3.5.1 Association of cortical myelin with demographic and clinical variables

The association of cortical myelin in the 33 selected parcels with demographic and clinical variables (see Table 1) was not statistically significant (Supplemental Table 4). Across all participants neither sex, IQ, nor MOODs-SR score were correlated with cortical myelin in selected parcels, nor did they interact with group (HC/UD) to predict cortical myelin. There was evidence for associations between age and cortical myelin in two regions (p-fdr<0.05), driven by a positive correlation in right LBelt, and a negative correlation in left a24pr. In individuals with UD, no clinical variable was correlated with cortical myelin in selected parcels. However, taking antidepressants was nominally associated (p<0.05 uncorrected) with increased cortical myelin in right OP1, left 24dv, and left p32, and decreased cortical myelin in right LBelt (Supplemental Table 4, Supplemental Figure 3).

#### 3.5.2 Association of LDA accuracy with demographic and clinical variables

Analyses found that no demographic or clinical variable was predictive of classification accuracy (Table 3). Similarly, within the UD participant group, no clinical or medication variable was associated with classification accuracy (Table 3).

**Table 3.**
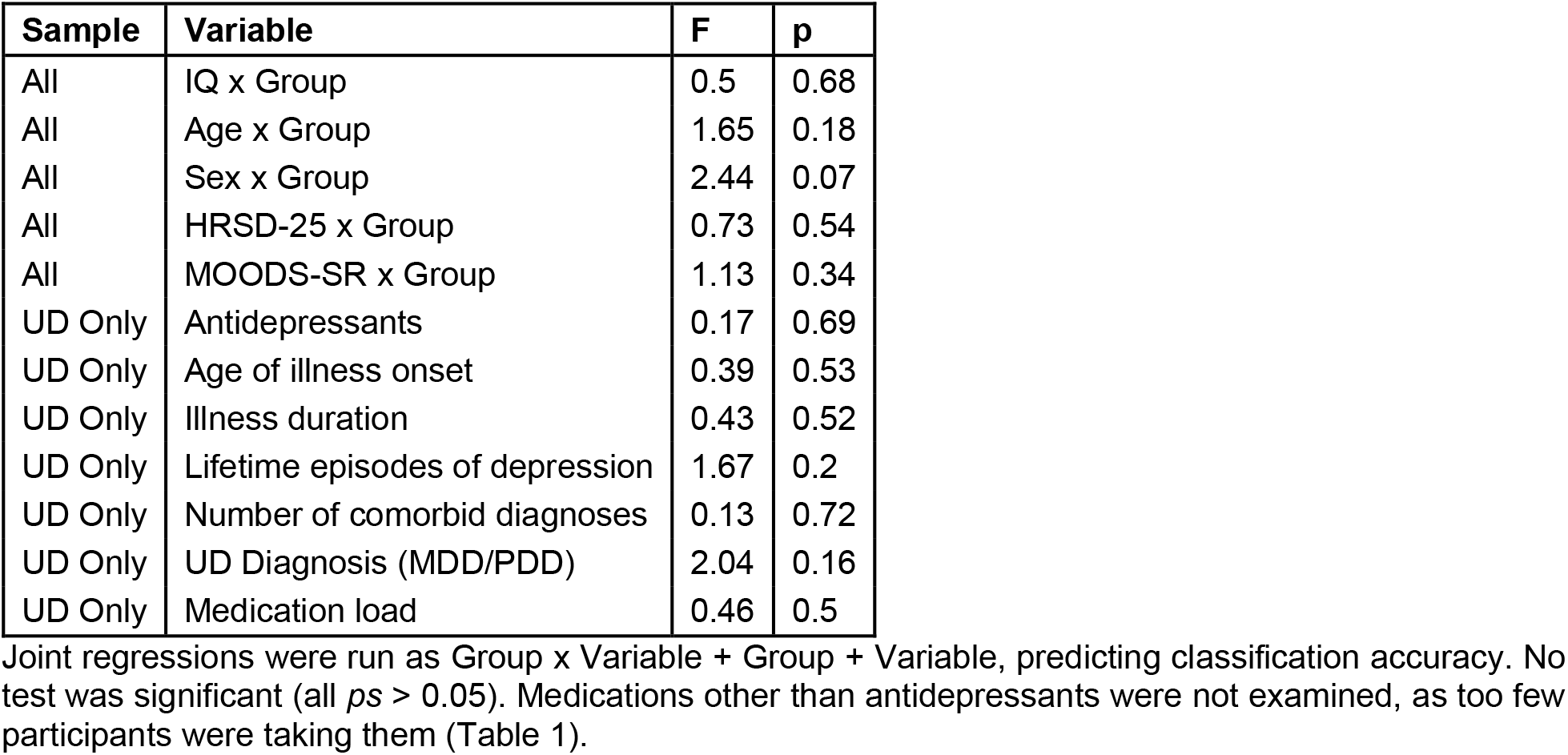
Association of demographic and clinical variables with classification accuracy

#### 3.5.3 Exploratory analysis of classification in a participant who converted from HC to UD

The LDA trained on the whole sample of 86 participants, with IQ and the 33 parcels identified in the previous analyses as predictors classified this participant as ‘UD’ both times: 12 months and 6 months before illness onset.

## 4 DISCUSSION

Cortical myelin distinguished healthy control participants (HC) from individuals with unipolar depression (UD) with 68% accuracy. The elastic net regression selected regions implicated in executive function and reward processing, as well as those involved in visual, auditory, and somatomotor processing. The pattern of cortical myelin distinguishing UD from HC was associated with a myelin reduction in some regions and increased myelin in other regions. These results demonstrate that aberrant levels of cortical myelin may be a biomarker of UD.

The 33 selected parcels included 13 parcels in regions important for executive functioning, cognitive control, and reward processing, including the anterior and posterior cingulate, orbitofrontal cortex, frontal operculum, and parietal cortex. There is abundant evidence that these processes are disrupted in depression (5,11). The present results converge with mounting evidence of disrupted myelination (28), thickness (12), connectivity (58), and activation (11) of these regions in those with depression. In addition to regions traditionally reported in neuroimaging studies of depression, we found that parcels located along the dorsal and ventral visual pathways, somatosensory and auditory processing regions and regions implicated in language also contributed to UD vs. HC classification. While depression is not typically considered a disorder of dysfunctional sensory processing, some studies report visual (59) and auditory (60) processing deficits in depression. In addition, somatosensory symptoms (e.g., motor retardation) are among the diagnostic criteria for this disorder and have been well described (61). Our results add to a growing body of literature documenting associations of depression with altered structure (12) and functional connectivity (62,63) in sensory regions. Broadly, our results contribute to the consensus that the neurobiology of regions that play important roles in cognitive processes and information transfer are disrupted in depression, which may contribute to the etiology of the disorder.

It is notable that in contrast to studies of the major white matter tracts, where depression is associated with lower integrity (15), we observed both decreased and increased cortical myelin in individuals with UD relative to HC. Additionally, the anterior cingulate, the superior parietal cortex, the retroinsular cortex, and visual processing regions included a combination of proximal regions with myelin decreases and increases. While the relationship between cortical myelin and functional activation or connectivity remains under-explored, these observations suggest that cortical myelin imbalance, rather than a uniform reduction, could drive some of the observed functional differences in depression, such as disrupted network integration (63,64) and patterns of both hypo-and hyper-connectivity (65,66).

Intriguingly, while classification accuracy was bimodal, in permutations participants were classified at chance (50%). This suggests that participants who were consistently misclassified differed from those who were correctly classified in their demographic or clinical features. However, *post-hoc* analyses did not reveal any associations between classification accuracy and demographic or clinical variables, thus suggesting that misclassification could reflect the presence of environmental or genetic risk factors that are not captured by clinical measures. These factors could alter the myelin content in selected regions thus creating predisposition to depression at the brain level. As a preliminary exploration of this idea, we explored the myelin patterns in a participant who experienced the onset of UD during the course of the study and was not used in primary analyses. This participant completed two MRI scans: at baseline and 6 months follow-up with both time points occurring prior to UD onset. Remarkably, this participant was classified as UD on both scans, despite not yet meeting criteria for a UD diagnosis at the time of scan. This preliminary result suggests that the pattern of cortical myelin in frontal, sensorimotor and extended visual cortices may be a biological risk marker predictive of UD diagnosis in the future. Further longitudinal studies are needed to test this hypothesis.

While the present results demonstrate that the pattern of cortical myelin is disrupted in UD, the cause of myelin disruption remains unknown. *Post-hoc* analyses suggest that myelin disruption is not related to medication use, lifetime depression severity, or illness duration. However, disrupted cortical myelin may reflect other risk factors for UD. For instance, sleep disturbance, which is a well-established risk factor and symptom of UD (67), was recently shown to correlate with cortical myelin in several of the same regions found in the present study, including the cingulate and middle temporal cortex (32). Stress is also associated with reduced white matter integrity in depression (15,68,69). After stress, remyelination can occur (70) thus resulting into altered patterns of myelination across the cortex (71). This ‘remyelination’ hypothesis could potentially explain our observations of both decreased and increased cortical myelin in unipolar depression.

Limitations of this work includes the need to replicate our findings in an independent sample. To partially address this limitation, our analyses used robust machine learning methods involving model testing using held-out samples. The major strength of this approach is that it helps to reduce model bias, which occurs when the same participants are used to train and test a model (72). It has been suggested that model performance with nested cross-validation is close to the accuracy that would be achieved on fully independent data (73). The second limitation concerns measurement noise due to susceptibility artifacts. Several regions with documented relevance to depression, including the bilateral hippocampus, entorhinal cortex, and posterior orbitofrontal cortex complex, were not included in the present analyses, as these regions showed an excess of between-person variability (see Supplemental Methods). Future research should explore the association of cortical myelin in these regions with unipolar depression.

In summary, cortical myelin can distinguish participants with UD from HC, even when clinical and demographic variables are not included in analyses. Regions that were most important for this classification include several that play key roles in reward and emotion processing as well as a host of regions important for sensory processing. This result highlights that the association of UD with sensory processing bears further investigation. Notably, UD was associated with both decreased and increased cortical myelin, suggesting that observations of reduced integrity of major white matter tracts in UD may not fully extend to the cortex. These results suggest that cortical myelin holds promise as a biomarker of unipolar depression and may be an early predictor of risk for this disorder.

## Supporting information

Primary Supplement

## Data Availability

Data will be made publicly available upon publication.

## ACKNOWLEDGMENTS

Funding Acknowledgements: This work was supported by a grant from the National Institute of Health R01MH114870 to A.M., and Y.O.H was supported by P41EB019936 to the Center for Reproducible Neuroimaging Computation (PI: Kennedy). D.A.A.B, Y.O.H., S.S., R.R, S.I., and A.M.: declare no conflict of interest. H.A.S: receives royalties from Wolters Kluwer, royalties and an editorial stipend from APA Press, and honorarium from Novus Medical Education.

The authors thank participants for taking part in this research study. We also thank Dr. Mary L. Phillips for fruitful discussions of the study design.

## AUTHORS CONTRIBUTION

D.B. – data quality assurance, analysis, visualization, and interpretation; drafted and critically evaluated the manuscript

Y.O.H. – curated data organization and analyses, drafted and critically evaluated the manuscript

S.S., R.R. – data acquisition and quality assurance; drafted and critically evaluated the manuscript

S.I. - curated data analyses, interpreted the data, critically evaluated the manuscript H.A.S., M.L.P. – curated participants’ recruitment, interpreted the data, critically evaluated the manuscript.

A.M. – obtained funding; designed the study; data acquisition, quality assurance, analysis and interpretation; drafted and critically evaluated the manuscript

All authors have approved the final version of the manuscript and agreed to be accountable for all aspects of this work.

