## Supplementary material for "Aberrant levels of cortical myelin distinguish individuals with unipolar depression from healthy controls": Primary Supplement

**Supplementary Information Table of Contents:**

Supplemental Methods: Pages 3-5

Supplemental Figures 1-3: Pages 6-8

Supplemental Tables 1-4: See Supplemental data file

**Supplemental Methods:**

*Subject-level quality control with mriqc*

Participants were excluded from the analyses if visual inspection identified the presence of gross motion or scanner-related artifacts in the T1w or T2w images, or in the *mriqc* background noise images, (e.g., ghosting, blurring, ringing, banding, etc.). The distribution of *mriqc* image quality metrics (IQMs) reflecting scan noise in our study were compared to the distribution of IQMs collected by the *mriqc* server from other studies and available via its API, from 1046 T1w and 619 T2w images collected using similar parameters (1). Scans with IQMs beyond the interquartile range of the *mriqc* API data (median +/- 1.5 x 75% quartile – 25% quartile) were flagged as potential outliers and were re-inspected. The distributions of the *mriqc* IQMs for the remaining participants were not different from the distribution of IQMs from the *mriqc* API (Supplemental Figures 1&2).

*Group-level preprocessing*

Data quality in some brain regions is worse than in the other regions due to susceptibility artifacts (2). This inconsistent quality of data may increase variability in myelin values across participants. The distribution of myelin values across all participants in each of the 360 regions was examined to identify regions whose variability was an outlier relative to variability in other regions. The coefficient of variation (sd/|mean|) was used to summarize the variability within each region. Rosner's test for outliers (3,4) identified 11 outlier parcels with excessively high variation (Supplemental Table 1) including parcels located in bilateral hippocampus, entorhinal cortex, presubiculum, piriform cortex, and posterior orbitofrontal cortex complex, and the right subgenual cingulate (bilateral H, EC, PreS, Pir, pOFC, and right 25).These regions, which are known to suffer from an excess of susceptibility artifacts (5,6), were removed from the analyses, leaving 349 parcels.

*Below is a step-by-step description of the nested-cross validation analysis.*

1. We identified all possible combinations of UD and HC participants to be held out in the nested analysis: 39 UD * 47 HC = 1833 combinations. This determined the number of loops of nested cross-validation.

2. In each loop of nested cross-validation, one UD/HC participant pair was set aside. The remaining 84 participants were used in the elastic net analysis and were then used to run the linear discriminant analysis (LDA). The UD/HC pair that was set aside was then used to test the LDA model as the final step in each loop. As there were 1833 unique UD/HC participant pairs, the nested analysis was run 1833 times, with 84 participants in the training data and a different UD and HC pair set aside as testing data. Each of the 1833 nested analyses was conducted in the following way:

    a. First, logistic elastic net was used to predict UD/HC status using cortical myelin values from n=349 parcels, age, IQ, and sex. Leave-one out cross validation (i.e., train the model on 83 participants, test on 1 participant, repeat 84 times) was used to identify the optimal $\lambda$ that corresponded to the minimal mean cross-validation error + 1 SE (7).

    b. Second, the optimal $\lambda$ was used to fit an elastic net model and select variables important for UD vs. HC classification.

c. Third, this set of variables was used to train the LDA model on the same 84 participants.

d. Fourth, the LDA model was then tested on the two participants (1 UD and 1 HC) that were held out in each nested cross-validation loop. If elastic net did not identify any variables beyond the model intercept, then both participants were recorded as misclassified.

3. As each participant was tested using LDA 84 times, participant-wise accuracy was computed as the mean of 84 LDA accuracies. Total model accuracy was computed as the average of the participant-wise accuracies. Model sensitivity and specificity were computed as the average of UD-only and HC-only participant accuracies, respectively.

4. The list of all variables selected by at least one elastic net model was generated. For each variable on this list, the proportion of the n=1883 models in which it was selected by the elastic-net regression was computed (i.e., variables selected in every model would be 100%, while variables that were selected in one model would be 0.0545%).

5. In order to identify the noise level for the frequency of variable selection as well as model fit, we repeated the same procedures described above while permuting UD/HC labels. In the permuted-labels analysis, each of 1833 loops was repeated 100 times for each set of 84 retained (training set) + 2 held-out (testing set) participants. To ensure unique randomization of UD/HC labels in each training set, a different random seed was used for each of 1833*100=183300 loops. True labels were kept for participants in the testing set. As UD/HC labels in the training sets were randomized, these classification results reflect model performance when myelin values do not carry useful information for distinguishing UD from HC (i.e., they reflect false-positive results and over-fitting). Accuracy and variable selection frequency were computed as described in the procedures for true-label analysis.

6. To identify the variables most strongly predictive of case/control status, and which are less likely to reflect noise, the variable selection frequency with true case labels was compared to the variable selection frequency with permuted case labels. The variable in the true-label model was retained if it was above selection frequency for this variable in the permuted-label model plus 3.5 IQR. For example, if a variable was selected in 75% of models using the true case labels, but only in 2% of models with the permuted labels, then that variable was retained.

*Supplemental References*


1. Beard E. MRIQCeption. 2020.
2. Olman CA, Davachi L, Inati S. Distortion and signal loss in medial temporal lobe. PLoS ONE. 2009;4(12).
3. Rosner B. On the detection of many outliers. Technometrics. 1975;
4. Millard SP. EnvStats, an RPackage for Environmental Statistics. In: Wiley StatsRef: Statistics Reference Online. 2014.
5. Deichmann R, Gottfried JA, Hutton C, Turner R. Optimized EPI for fMRI studies of the orbitofrontal cortex. NeuroImage. 2003;19(2):430–41.
6. Weiskopf N, Hutton C, Josephs O, Deichmann R. Optimal EPI parameters for reduction of susceptibility-induced BOLD sensitivity losses: A whole-brain analysis at 3 T and 1.5 T. NeuroImage. 2006;33(2):493–504.
7. Friedman J, Hastie T, Tibshirani R. Regularization paths for generalized linear models via coordinate descent. Journal of Statistical Software. 2010;

Supplemental Figure 1. Comparison of T1 mriqc values between study data and API data

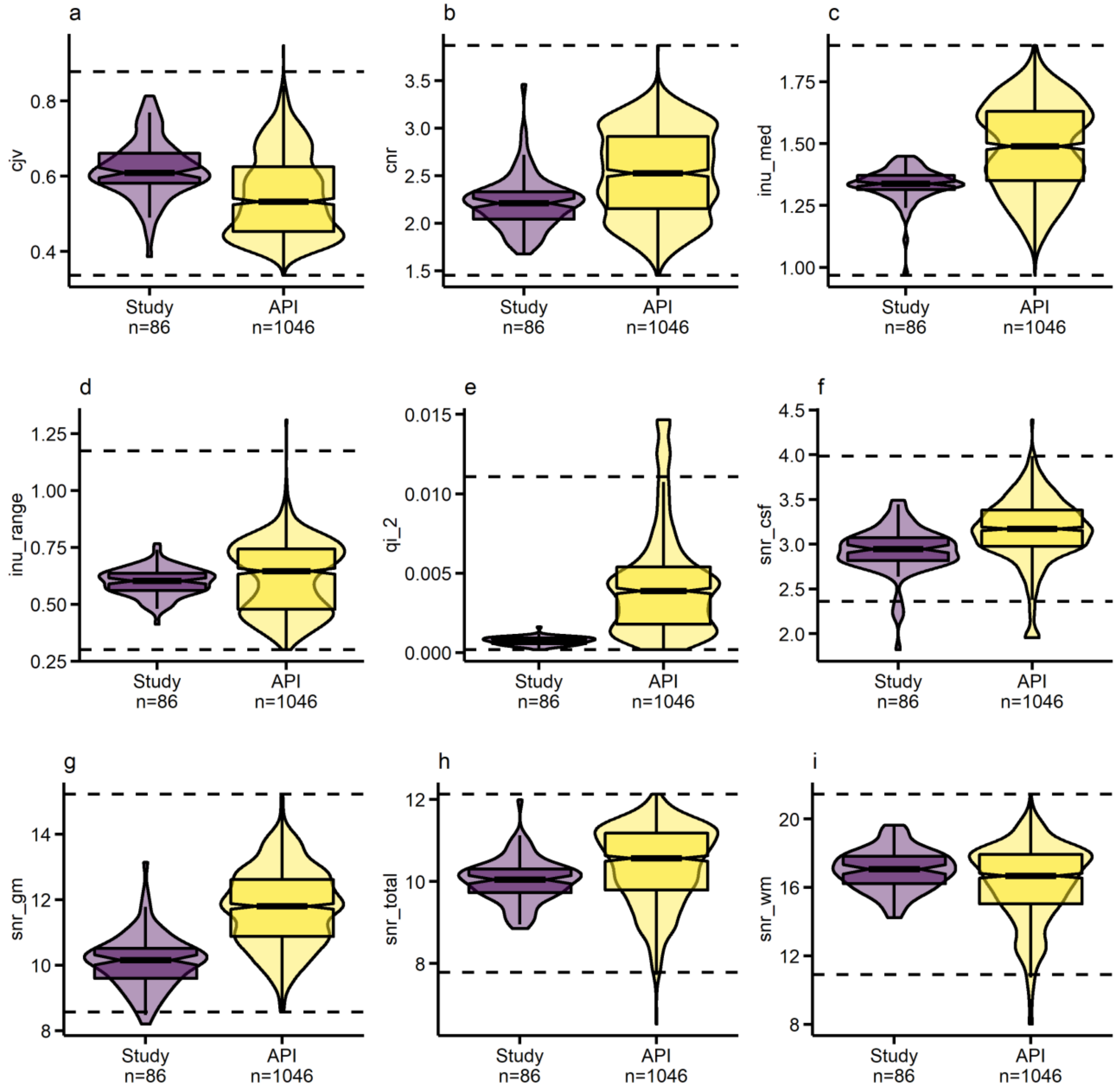


**a**) Coefficient of joint variation between white matter and gray matter. Higher values indicate more head motion and/or intensity non-uniformity artifacts. **b**) Contrast-to-noise ratio, reflecting separation between GM & WM. Higher values indicate higher quality. **c**) Intensity non-uniformity (bias field) median. Values closer to 1 indicate higher quality; further from zero indicate greater RF field inhomogeneity. **d**) Intensity non-uniformity (bias field) range. Values closer to 1 indicate higher quality; further from zero indicate greater RF field inhomogeneity. **e**) Mortamet's quality index 2. A quality index accounting for effects of both clustered and subtle artifacts in the air background. Higher values indicate lower quality. **f**) Signal-to-noise ratio within the CSF mask. Higher values indicate higher quality. **g**) Signal-to-noise ratio within the grey matter mask. Higher values indicate higher quality. **h**) Signal-to-noise ratio within the total mask. Higher values indicate higher quality. **i**) Signal-to-noise ratio within the white matter mask. Higher values indicate higher quality.

Supplemental Figure 2. Comparison of T2 mriqc values between study data and API data

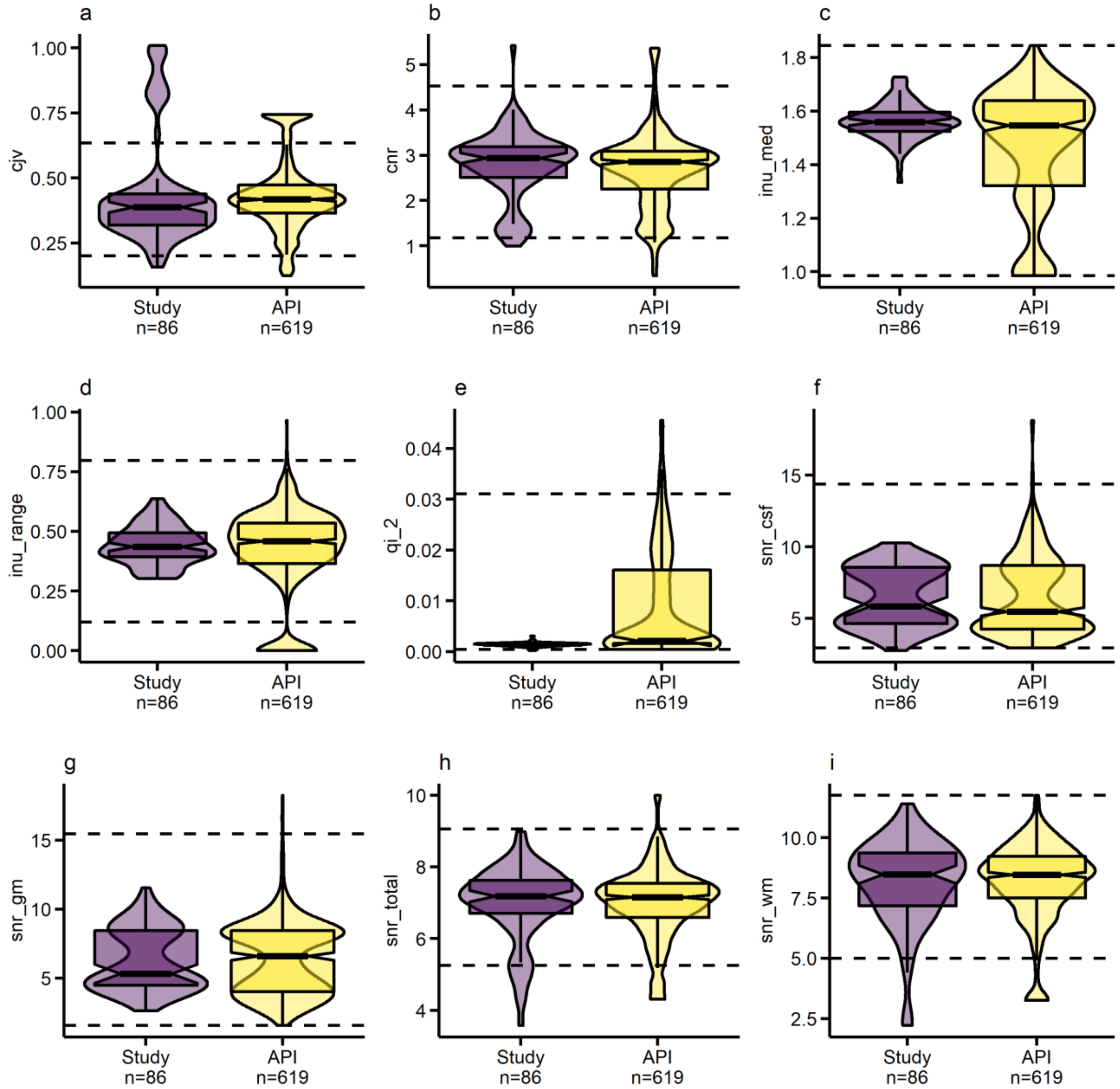


**a**) Coefficient of joint variation between white matter and gray matter. Higher values indicate more head motion and/or intensity non-uniformity artifacts. **b**) Contrast-to-noise ratio, reflecting separation between GM & WM. Higher values indicate higher quality. **c**) Intensity non-uniformity (bias field) median. Values closer to 1 indicate higher quality; further from zero indicate greater RF field inhomogeneity. **d**) Intensity non-uniformity (bias field) range. Values closer to 1 indicate higher quality; further from zero indicate greater RF field inhomogeneity. **e**) Mortamet's quality index 2. A quality index accounting for effects of both clustered and subtle artifacts in the air background. Higher values indicate lower quality. **f**) Signal-to-noise ratio within the CSF mask. Higher values indicate higher quality. **g**) Signal-to-noise ratio within the grey matter mask. Higher values indicate higher quality. **h**) Signal-to-noise ratio within the total mask. Higher values indicate higher quality. **i**) Signal-to-noise ratio within the white matter mask. Higher values indicate higher quality.

Supplemental Figure 3. Association of antidepressant medication with cortical myelin

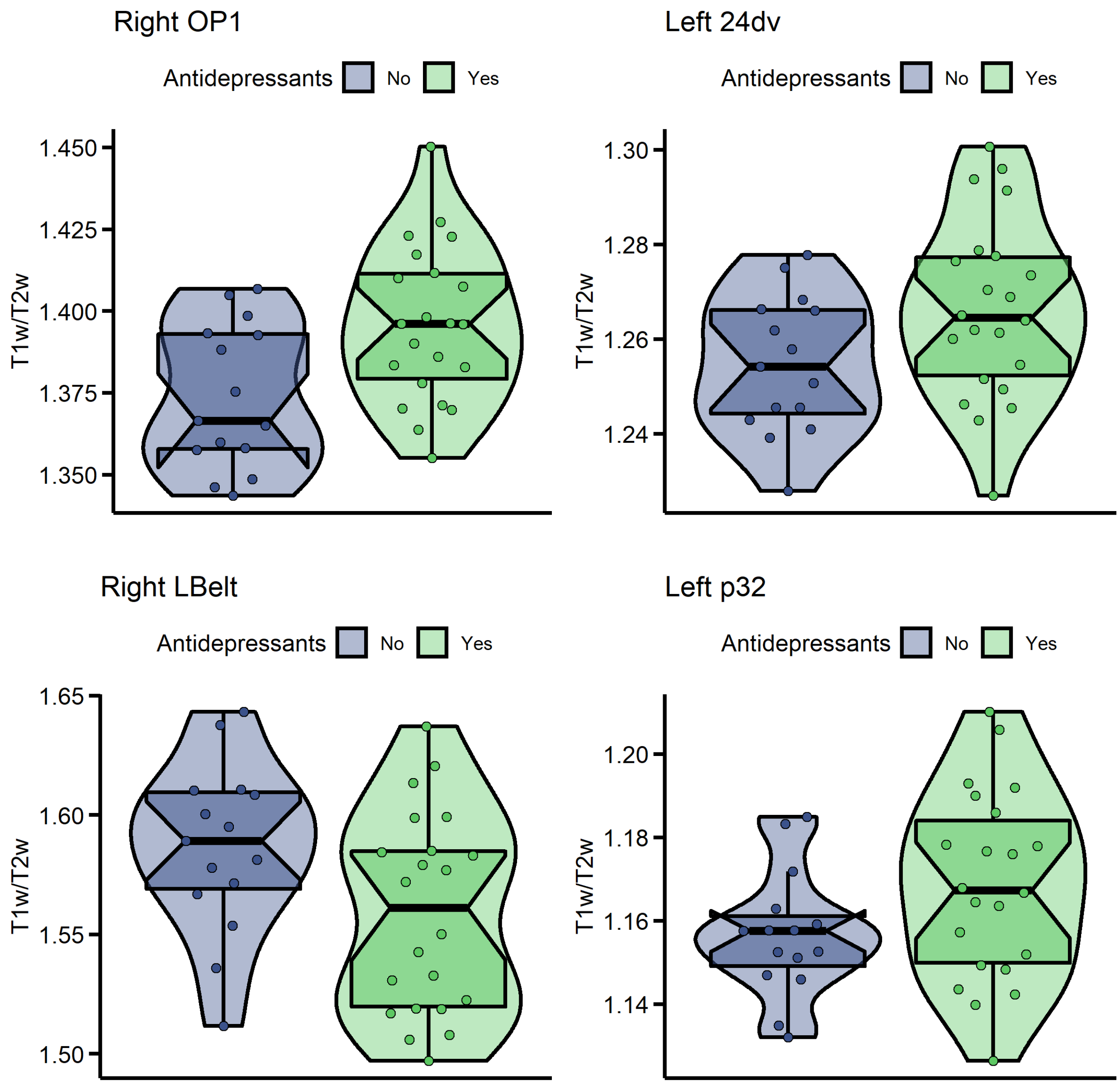


In participants with UD (n=39), antidepressant medication (n=22 were taking antidepressants, n=17 were not) was nominally associated (p<0.05 uncorrected) with cortical myelin in 4 of the 33 selected parcels.
